# Assessing the Quality of Electronic Health Record Data and the Claims Linked Data for Target Trial Emulation Studies

**DOI:** 10.64898/2025.12.22.25342844

**Authors:** Yao An Lee, Ying Lu, Earl J Morris, Xing He, Almut G Winterstein, Carl Henriksen, Jiang Bian, Jingchuan Guo

**Author notes:** Corresponding Author: Jingchuan Guo, MD, PhD, Department of Pharmacy Practice, Purdue University College of Pharmacy Address: 640 Eskenazi Ave, Indianapolis, IN 46202. Funding support: NIH/NIA R01AG089445; NIH/NIDDK R01DK133465.

## Abstract

**Objectives:** To evaluate whether a EHR cohort, alone and linked to Medicare claims, has sufficient data quality to support design elements required for target trial emulation, using type 2 diabetes (T2D) as a case example.

**Materials and Methods:** We constructed annual University of Florida Health EHR–Medicare linked cohorts of patients ≥ 65 years with T2D from 2013 to 2020. Using Medicare claims as the reference, we assessed EHR data quality for target trial emulation-relevant elements across completeness, accuracy, plausibility, and concordance, spanning target trial components (eligibility, exposure/new-user ascertainment, baseline covariates, outcomes, and follow-up). Data quality was compared across EHR-only, claims-only, and EHR–claims linked data.

**Results:** The mean annual EHR–Medicare linked cohort included 12,895 patients (mean age 74.9 years; 58.0% female). Demographics were complete and highly accurate. In the EHR-only cohort, completeness ranged 34.1–78.4% for conditions and 53.7–63.4% for glucose lowering drugs (GLDs). Accuracy was high for prevalent conditions and GLD use but low for incident measures. Plausible values were common (>98.5%), and HbA1c - T2D concordance was strong (98.6%). Linking EHR and claims substantially improved completeness and accuracy, especially for encounters, mortality, incident diagnoses, and medications.

**Discussion:** The linked dataset addressed major limitations of EHR-only data and provided enhanced granularity compared to claims alone, offering a comprehensive resource for real-world target trial emulation research.

**Conclusion:** EHRs offer valuable clinical details but face data quality challenges. Robust quality assurance strategies and linkage with external data are essential to strengthen real-world evidence and support target trial emulation.

**Lay Summary:** We evaluated whether a University of Florida Health electronic health record (EHR) cohort—alone and when linked to Medicare claims—has sufficient data quality to support “target trial emulation,” a common approach for using real-world data to study medication effects when randomized trials are not feasible. We studied adults aged 65 years and older with type 2 diabetes from 2013–2020 and assessed four practical dimensions of data quality: completeness (how often key information is captured), accuracy (agreement with Medicare for billing-derived elements), plausibility (whether recorded values are clinically reasonable), and concordance (internal consistency between related EHR elements). Demographic fields were highly complete and accurate, and most lab and vital sign values were biologically plausible, supporting the reliability of core EHR clinical measurements. However, the EHR alone missed a substantial share of encounters, deaths, incident diagnoses, and medication initiation events that appeared in Medicare, reflecting care received outside a single health system. Linking EHR with Medicare substantially improved capture of these cross-setting events while preserving EHR-only clinical details (e.g., HbA1c and BMI), yielding a more robust dataset for real-world target trial emulation research.

## Background

The growing importance of real-world evidence (RWE) in healthcare research, clinical decision-making, and policy development has highlighted the need for robust real-world data (RWD) sources[1–3]. Among various RWD sources, including insurance claims databases, registries, and patient-reported outcomes, electronic health records (EHRs) have emerged as one of the most widely used sources due to their widespread adoption and rich clinical detail[4,5]. As of 2021, over 75% of office-based physicians and nearly all non-federal acute care hospitals in the U.S. utilized certified EHR systems[6]. For chronic conditions such as type 2 diabetes (T2D), a progressive metabolic disorder affecting over 34 million Americans[7], EHRs provide detailed clinical information, including laboratory values (e.g., hemoglobin A1c [HbA1c]), clinical observations (e.g., body mass index [BMI]), prescribed medications, comorbidities, and complications, making them valuable for tracking disease progression and care quality over time[8]. Despite these strengths, EHRs were primarily designed for supporting clinical care and administrative workflows rather than research. Therefore, they often present notable data collection challenges, including incomplete documentation, inconsistent coding, and limited interoperability[9–11]. These challenges can compromise data reliability and data validity, especially for chronic conditions like T2D that require continuous, multisite care coordination.

In contrast, insurance claims data offer more complete documentation of reimbursed healthcare services across providers and care settings, making them particularly useful for tracking healthcare utilization patterns, diagnoses, procedures, and mortality among older adults[12–14]. Insurance claims data lack the clinical granularity of EHRs, such as lab results and vital signs. Nevertheless, their standardization, longitudinal coverage, and national scope make them a valuable benchmark for evaluating the completeness, accuracy, and representativeness of EHR-based data. Integrating EHRs with Medicare claims can therefore provide a more comprehensive and accurate picture of patient care, combining detailed clinical insight with broad healthcare utilization patterns[15,16]. Although linked EHR-Claims data have gained increasing attention for clinical research, few studies have systematically assessed the quality of such linked datasets across multiple domains.

For target trial emulation studies using RWD to evaluate drug benefits and risks, complete and valid information on drug use (e.g., new users vs. prevalent users) and condition information (e.g., incident vs. prevalent cases) is critical for causal inference between the drug of interest and the clinical outcome of interest. However, it remains unclear whether single-site EHR data alone can consistently support these target trial emulation-critical elements—particularly treatment initiation, incident outcome capture, baseline covariates for confounding control, and follow-up definition—and how much linkage to claims improves these limitations. Therefore, this study aims to fill that gap by systematically evaluating the quality of a cohort linking University of Florida (UF) Health EHR data with Medicare claims, using T2D target trial emulation studies as a case example. We selected T2D because it is highly prevalent among older adults and requires longitudinal medication and outcome capture, and we used Medicare claims because they provide broad, standardized longitudinal data for adults ≥65 years and are the primary claims source available for linkage in our institute.

## Material and methods

### Data sources and population

This retrospective cohort study evaluated the quality of EHR data from the UF Health system by linking it with Medicare claims data.

UF Health EHR data were obtained from the UF Health Integrated Data Repository (IDR), an enterprise data warehouse that consolidates patient-level data from various clinical and administrative systems across the UF Health system. UF Health is a large academic healthcare network comprising 12 hospitals and hundreds of outpatient clinics across Gainesville, Jacksonville, and Central Florida, collectively serving over 3 million inpatient and outpatient visits annually[17]. Medicare claims data were obtained from the Centers for Medicare & Medicaid Services (CMS), a federal health insurance program covering adults aged ≥65 years and individuals with qualifying disabilities. We restricted the cohort to inpatient (Part A), outpatient (Part B), and prescription drug (Part D) claims because Medicare Advantage claims are not consistently complete in our data environment due to private plan reporting. We required continuous Medicare enrollment during each measurement window to ensure complete capture of healthcare utilization, diagnoses, and medication claims; beneficiaries with partial-year enrollment were excluded.

The study population included older adults aged 65 years with T2D, identified in Medicare claims between January 1, 2013, and December 31, 2020, using validated International Classification of Diseases (ICD) codes. Individuals with type 1 diabetes (T1D) were excluded (**Table S1).** T1D and T2D were defined using CCW-based identification criteria applied to Medicare claims during the study period. The UF Health EHR-Medicare linked cohort consisted of individuals who received care within the UF Health system and were successfully matched with Medicare data, with Medicare claims capturing billed services both within UF Health and at external providers/settings during the study period. Linkage was performed using a deterministic matching algorithm based on Social Security Number (SSN), date of birth (DOB), and sex (**Figure S1**). Specifically, UF Health provided a secure crosswalk between UF Health patient identifiers and SSNs to the CMS contractor (GDIT), which crosswalked SSNs to Medicare beneficiary identifiers (BENE_IDs) and returned an anonymized UF Health ID–to–BENE_ID linkage file. Linkage was performed at the person level (not encounter/visit level). All personally identifiable information was removed following linkage to protect patient privacy. For the linked cohort, we separately extracted key variables from both UF Health EHR data and Medicare claims and evaluated the quality of EHR data by benchmarking it against corresponding Medicare data. We used Medicare fee-for-service claims as a pragmatic reference standard for billing-derived domains (e.g., encounters, diagnoses/procedures, and dispensed medications), because claim elements required for reimbursement tend to have higher data fidelity and provide longitudinal capture of billed care beyond a single health system**[18,19]**; therefore, we benchmarked corresponding EHR-derived variables against claims to assess EHR data quality and the incremental value of linkage. The study was approved by the Institutional Review Board (IRB) at the University of Florida.

### Variables assessed

We examined a wide range of variables across six domains: demographics, mortality, clinical measurements (i.e., clinical observations and laboratory values), healthcare utilization, clinical conditions, and glucose-lowering drug (GLD) use. The variable selection is based on the target trial emulation framework (**Table S2**)[20,21]. To align with target trial specification, we grouped variables by core target trial emulation components. Eligibility was assessed via T2D identification; treatment/new-user feasibility via GLD initiation using a look-back period; confounding control via baseline demographics, comorbidities, and key clinical measures; and outcomes/follow-up via mortality, incident diagnoses, and encounters. This mapping ensures each data quality metric informs a specific component required for emulation.

Demographic variables included age, sex, and race/ethnicity (categorized as Non-Hispanic White [NHW], Non-Hispanic Black [NHB], Hispanic, other, or unknown). Mortality data were captured based on documented dates of death. Clinical measurements included HbA1c and BMI, with BMI values derived from either direct documentation or calculation based on height and weight data. HbA1c was analyzed from EHR laboratory results and reported as percent (%), and BMI was reported in kg/m²; Medicare claims do not contain lab result and BMI values in our dataset. Healthcare utilization was measured using the number of encounters per year, defined as inpatient admissions and outpatient encounters (including emergency department and ambulatory visits) in UF Health EHRs; in Medicare claims, inpatient encounters were derived from Part A and outpatient/physician services from Part B. Clinical conditions frequently comorbid with T2D in older adults who were examined, were defined using ICD-9-CM codes prior to 2015 Q4 and ICD-10-CM codes from 2015 Q4 onward (**Table S1**), including chronic kidney disease (CKD), heart failure (HF), hypertension (HTN), depression, stroke or transient ischemic attack (Stroke/TIA), acute myocardial infarction (AMI), and Alzheimer’s disease and related dementias (ADRD). GLD use was assessed for six key drug classes (**Table S3**): Metformin (MET), glucagon-like peptide-1 receptor agonists (GLP-1RAs), sodium-glucose cotransporter 2 inhibitors (SGLT2is), dipeptidyl peptidase-4 inhibitors (DPP4is), sulfonylureas (SUs), and thiazolidinediones (TZDs). Medication use was defined by the presence of standardized medication codes (RxNorm in EHR; NDC/Part D pharmacy claims), and dose or quantity information was not incorporated. Incident condition and incident GLD use were defined as having no record in the applicable look-back window. Incident condition definitions followed CCW algorithms[22] with a condition-specific look-back windows (**Table S1**); incident GLD use was defined using a 1-year look-back window (no record in the prior year), applied consistently in both UF Health EHR and Medicare claims. Each variable was extracted separately from the UF Health EHR and Medicare claims to support cross-source comparisons for data quality evaluation.

### Data quality assessment

We assessed the quality of UF Health EHR data within the linked EHR–Medicare cohort using four key dimensions - completeness, accuracy, plausibility, and concordance - adapted from the established framework by Weiskopf et al[23]. All metrics were assessed on an annual basis to capture temporal trends and to identify potential improvements or declines in EHR data quality over time, with annual rates calculated using the unique number of eligible patients contributing a full year of observation in each calendar year. We further calculated the mean and standard deviation (SD) across 2013-2020 years.

Completeness refers to the presence of expected data elements in the EHR, using claims data as the gold standard. For each domain—demographics, mortality, healthcare encounters, clinical conditions, and GLD use—we assessed completeness as EHR capture among linked patients with corresponding information captured in Medicare. For each domain, we defined “captured” in a source as follows: for event-based domains (lab tests, clinical conditions, and GLD use), having ≥1 qualifying record during the study window; for demographics and healthcare encounters (e.g., sex, race), having a non-missing recorded value. Completeness was then calculated as the proportion of patients with the element captured in Medicare who also had the element captured in the EHR:

***Completeness rate = Number of patients with element captured in EHR / Number of patients with element captured in Medicare***

Accuracy reflects how well EHR-recorded information matches Medicare claims. We measured accuracy as the patient-level proportion of agreement between EHR and Medicare among patients with the element captured in the EHR. For event-based domains (e.g., clinical conditions and GLD use), “captured” was defined as having ≥1 qualifying record during the study window; for demographics (e.g., sex, race), “captured” required a non-missing value and “matched” required the same value in Medicare. The proportion of accuracy was calculated as:

***Accuracy rate = Number of patients whose EHR information matched Medicare / Number of patients with the element captured in EHR***

Plausibility evaluates whether observed values documented in EHR fall within clinically reasonable ranges. Plausible entries were flagged if they met predefined thresholds: HbA1c value > 2.5% and < 25%[24], BMI > 12 and < 70 kg/m^2^[25], and encounter dates preceding birth or following death. The proportion of plausibility values was calculated as:

***Plausibility rate = Number of plausible values in EHR / Total number of values assessed in EHR***

Concordance assesses the internal consistency of related variables within the UF Health EHR data. Specifically, we examined the agreement between T2D diagnosis and laboratory evidence of T2D through HbA1c testing. For each calendar year, we identified patients with at least one HbA1c test value of ≥6.5%, the diagnostic threshold for diabetes according to clinical guidelines[26]. Among this group, we calculated the proportion of patients who also had a recorded T2D diagnosis in the EHR during the same year. Among the patients with at least one HbA1c test value of ≥6.5%, the concordance rate was calculated as:

***Concordance rate = Number of patients with T2D diagnosis in EHR / Total number of patients with at least one HbA1c test value of*** ≥***6.5% in EHR***

We also compared data quality for the T2D cohort across EHR-only, Claims-only, and EHR–Claims linked data to address the strengths and limitations of each data source. This comparison was intended to illustrate how reliance on a single source can yield different capture and agreement for TTE-relevant elements, and how linkage leverages the complementary strengths of EHR clinical detail and claims-based cross-setting ascertainment. Within the linked cohort, ‘EHR-only’ uses UF Health EHR records only and ‘Claims-only’ uses Medicare claims records only, each restricted to the same linked beneficiaries.

In addition, we assessed the magnitude of representativeness of the UF EHR-Medicare linked dataset vs. the T2D populations in the national Medicare claims data. This representativeness assessment was intended to contextualize the transportability of single-site EHR findings and to identify cohort differences that could influence downstream target trial emulation analyses. Specifically, we compared the characteristics of EHR data to those of the broader Medicare population with T2D at both the state (Florida) and national levels. The comparison focused on five domains: demographics, mortality, HbA1c testing, prevalence and incidence of clinical conditions, and GLD use. For each calendar year, we calculated the proportion of individuals with each characteristic in both the linked cohort and the national Medicare population. Differences between groups were evaluated using standardized mean differences (SMDs) and statistical tests (chi-square tests for categorical variables and t-tests for continuous variables). An absolute SMD of less than 0.10 was interpreted as a negligible difference, and a p-value < 0.05 was considered statistically significant.

## Results

### EHR and Medicare claims linked T2D cohort

The size of the UF Health EHR–Medicare linked cohort with T2D varied annually due to variations in healthcare utilization within the UF Health system and dynamic enrollment in Medicare. Between 2013 and 2020, the annual cohort included 12,895 patients with a mean age of 74.9 years. Women consistently comprised the majority, accounting for 58.0% of the cohort on average. NHW individuals represented the largest racial/ethnic group, accounting for 64.88% of the cohort (**Table S4**).

### Data quality assessment of EHR data

#### Completeness of EHR data

Detailed completeness metrics are summarized in **Table 1**. Demographic information was fully complete, with no missing data for DOB, sex, or race/ethnicity in the EHR. However, EHR-recorded deaths accounted for 68.2% (SD: 6.51) of total deaths observed in Medicare data. On average, EHRs captured 38.3% (SD: 2.56) of all healthcare encounters documented in Medicare claims, including 41.8% (SD: 5.00) of inpatient and 37.3% (SD: 2.48) of outpatient visits. HbA1c testing documented in the EHR accounted for 53.6% (SD: 4.53) of patients with tests recorded in Medicare claims, while BMI measurements were available for 82.4% of the cohort (SD: 2.42).

**Table 1.** Completeness of UF Health EHR data compared to Medicare claims in the UF Health EHR-Medicare linked T2D cohort (2013-2020)

| Characteristic | 2013 | 2014 | 2015 | 2016 | 2017 | 2018 | 2019 | 2020 | Mean (SD) |
| --- | --- | --- | --- | --- | --- | --- | --- | --- | --- |
| **Demographics (%)** |  |  |  |  |  |  |  |  |  |
| DOB | 100.00 | 100.00 | 100.00 | 100.00 | 100.00 | 100.00 | 100.00 | 100.00 | 100.00 (0.00) |
| Sex | 100.00 | 100.00 | 100.00 | 100.00 | 100.00 | 100.00 | 100.00 | 100.00 | 100.00 (0.00) |
| Race / Ethnicity | 100.00 | 100.00 | 100.00 | 100.00 | 100.00 | 100.00 | 100.00 | 100.00 | 100.00 (0.00) |
| **Mortality (%)** |  |  |  |  |  |  |  |  |  |
| Date of Death | 71.18 | 68.44 | 65.35 | 56.74 | 63.14 | 69.10 | 77.09 | 74.73 | 68.22 (6.51) |
| **Encounter (%)** |  |  |  |  |  |  |  |  |  |
| Overall Encounter | 36.36 | 39.6 | 41.61 | 35.09 | 36.69 | 36.25 | 39.49 | 41.60 | 38.34 (2.56) |
| Inpatient Encounter | 32.02 | 49.35 | 46.16 | 40.55 | 41.09 | 40.68 | 41.91 | 42.35 | 41.76 (5.00) |
| Outpatient Encounter | 36.09 | 38.69 | 40.38 | 34.02 | 35.63 | 35.00 | 38.24 | 40.52 | 37.32 (2.48) |
| **Lab test (%)** |  |  |  |  |  |  |  |  |  |
| HbA1c Test | 50.19 | 58.81 | 62.59 | 51.26 | 52.18 | 50.95 | 52.28 | 50.78 | 53.63 (4.53) |
| **Vital sign (%)** |  |  |  |  |  |  |  |  |  |
| BMI | 82.50 | 86.58 | 83.06 | 80.18 | 82.45 | 82.82 | 83.03 | 78.20 | 82.35 (2.42) |
| **Clinical conditions (%)** |  |  |  |  |  |  |  |  |  |
| CKD | 37.76 | 44.70 | 47.30 | 41.58 | 41.41 | 42.11 | 44.54 | 44.70 | 43.00 (2.92) |
| HF | 28.71 | 38.30 | 43.38 | 43.81 | 46.92 | 49.07 | 52.88 | 54.41 | 44.69 (8.32) |
| HTN | 67.86 | 76.33 | 78.22 | 77.47 | 78.79 | 80.55 | 83.19 | 84.51 | 78.37 (5.09) |
| T2D | 59.32 | 65.84 | 66.78 | 65.16 | 66.20 | 66.08 | 68.50 | 68.59 | 65.81 (2.90) |
| Depression | 29.34 | 39.43 | 44.19 | 43.42 | 46.24 | 47.78 | 51.71 | 51.92 | 44.25 (7.34) |
| Stroke / TIA | 36.08 | 46.07 | 51.18 | 50.35 | 54.00 | 55.98 | 58.79 | 61.20 | 51.71 (7.95) |
| AMI | 36.08 | 52.97 | 59.43 | 56.96 | 61.41 | 74.44 | 82.88 | 86.38 | 63.82 (16.67) |
| AD | 21.45 | 29.13 | 30.16 | 33.82 | 37.63 | 37.00 | 41.34 | 42.48 | 34.13 (7.01) |
| **GLD use (%)** |  |  |  |  |  |  |  |  |  |
| MET | 42.69 | 61.37 | 65.86 | 60.61 | 59.71 | 56.31 | 53.71 | 52.00 | 56.53 (7.14) |
| GLP-1RA | 38.52 | 51.22 | 61.33 | 52.74 | 60.87 | 63.45 | 64.99 | 64.48 | 57.20 (9.16) |
| SGLT2i | 0.00 | 73.47 | 61.84 | 57.59 | 49.63 | 65.64 | 61.34 | 59.67 | 53.65 (27.71) |
| DPP4i | 49.04 | 67.69 | 69.53 | 66.93 | 67.31 | 65.31 | 61.31 | 59.93 | 63.38 (6.65) |
| SU | 41.09 | 60.46 | 63.54 | 58.09 | 57.59 | 54.25 | 52.81 | 49.28 | 54.64 (7.07) |
| TZD | 45.63 | 63.07 | 66.67 | 60.05 | 59.27 | 57.47 | 57.01 | 54.88 | 58.01 (6.22) |

The completeness of clinical conditions in the EHR varied by condition (**Figure 1-A**). HTN had the highest completeness at 78.4% (SD: 5.09), followed by T2D at 65.8% (SD: 2.90), while AD had the least completeness at 34.1% (SD: 7.01). Similarly, the completeness of GLD documentation varied by drug classes and year (**Figure 1-B**). Among newer agents, DPP4i showed the highest completeness, averaging 63.4% (SD: 6.65), followed by GLP-1RAs at 57.2% (SD: 9.16), and SGLT2i at 53.7% (SD: 27.71). Among traditional GLDs, MET showed a mean completeness rate of 56.5% (SD: 7.14), SU of 54.6% (SD: 7.07), and TZD of 58.0% (SD: 6.22).

**Figure 1.**
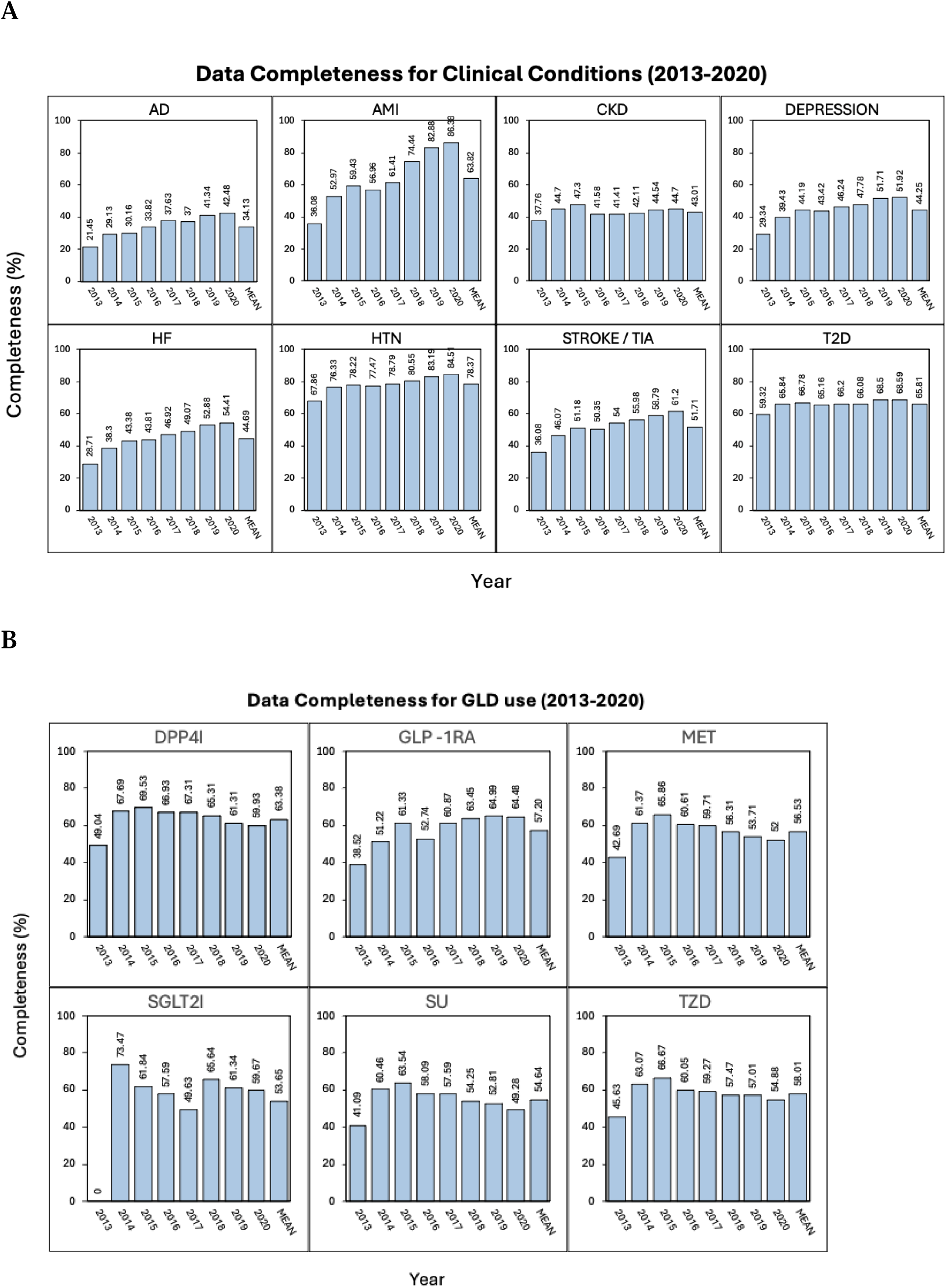
Completeness of UF Health EHR data compared to Medicare claims for key clinical conditions (A) and GLD use (B) in the UF Health EHR-Medicare linked T2D cohort (2013–2020). The proportion of Completeness values was calculated as: ***Completeness rate = Number of patients with element captured in EHR / Number of patients with element captured in Medicare***

#### Accuracy of EHR data

Detailed accuracy metrics are presented in **Table 2**. Overall, demographic data in the EHR showed high accuracy when compared with Medicare claims. Sex matched perfectly across all years, with no discrepancies. High accuracies were observed for DOB at 99.3% (SD: 0.20) and race/ethnicity at 92.6% (SD: 0.20), while mortality data had a moderate accuracy rate of 96.7% (SD: 0.85).

**Table 2.**
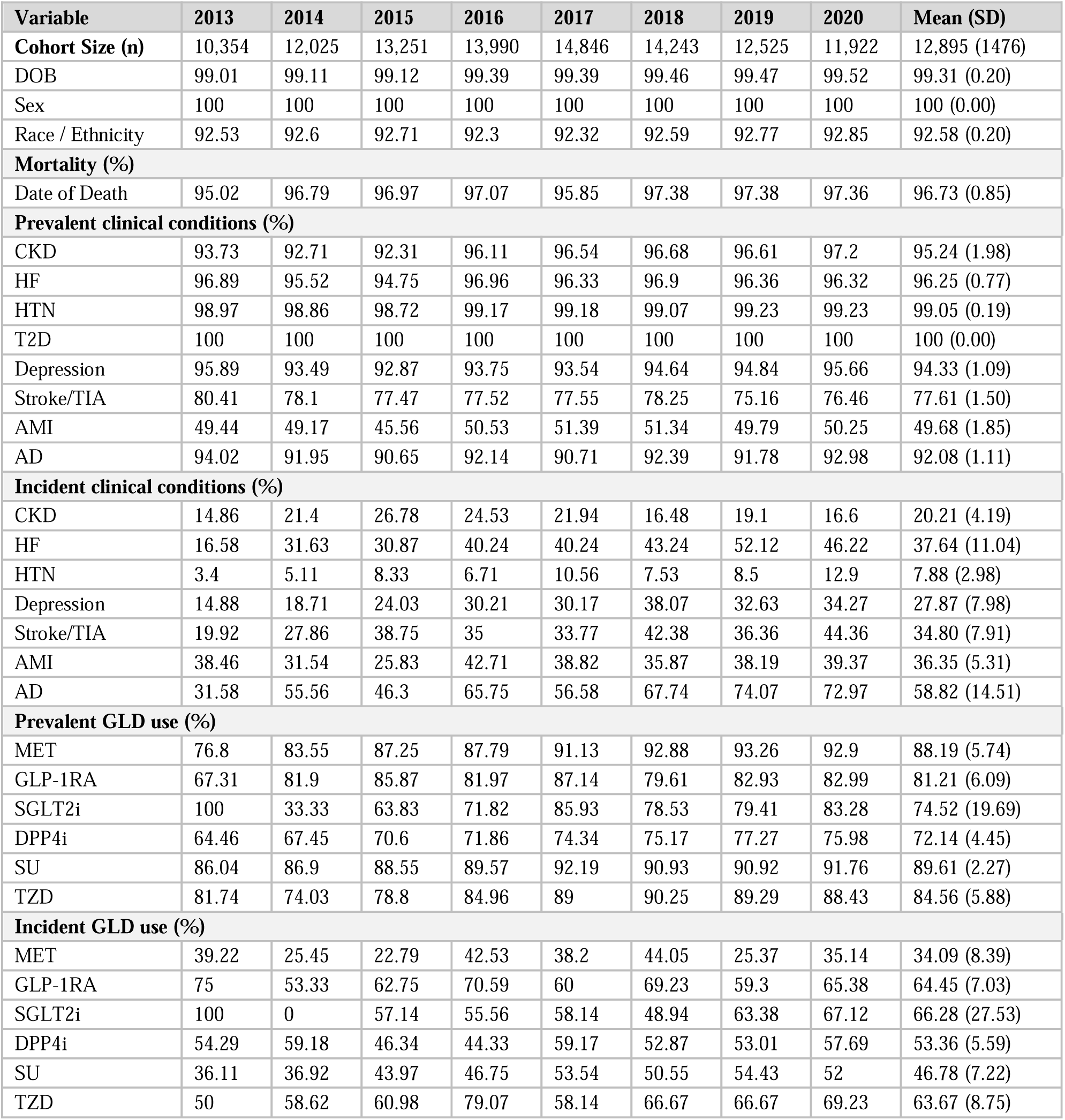
Accuracy of UF Health EHR data compared to Medicare claims in the UF Health EHR-Medicare linked T2D cohort (2013-2020)

The accuracy of clinical condition prevalence varied by diagnosis (**Figure 2-A**). T2D showed perfect alignment with Medicare claims (100.0% accuracy), and other common conditions such as HTN (99.05%, SD: 0.19), CKD (95.2% SD: 1.98), HF (96.2%, SD: 0.77), and depression (94.33%, SD: 1.09) also showed relatively high accuracy. However, more complex or acute conditions like stroke/TIA (78.6%, SD: 1.50) and AMI (49.7%, SD: 1.85) exhibited substantially higher discrepancies. Although accuracy trends improved over time for many conditions, AMI accuracy remained low (<55%) throughout the study period. In contrast, EHR capture of incident conditions (new diagnoses compared to the prior year) show consistently low accuracy (**Figure 2-B**). HTN had the lowest accuracy rate at 7.9% (SD: 2.98), followed by CKD (20.2%), depression (27.9%, SD: 7.98), stroke/TIA (34.8%, SD: 7.91), AMI (36.3%, SD: 5.31), HF (37.6%, SD: 11.04), and AD (58.8%, SD: 14.51).

**Figure 2.**
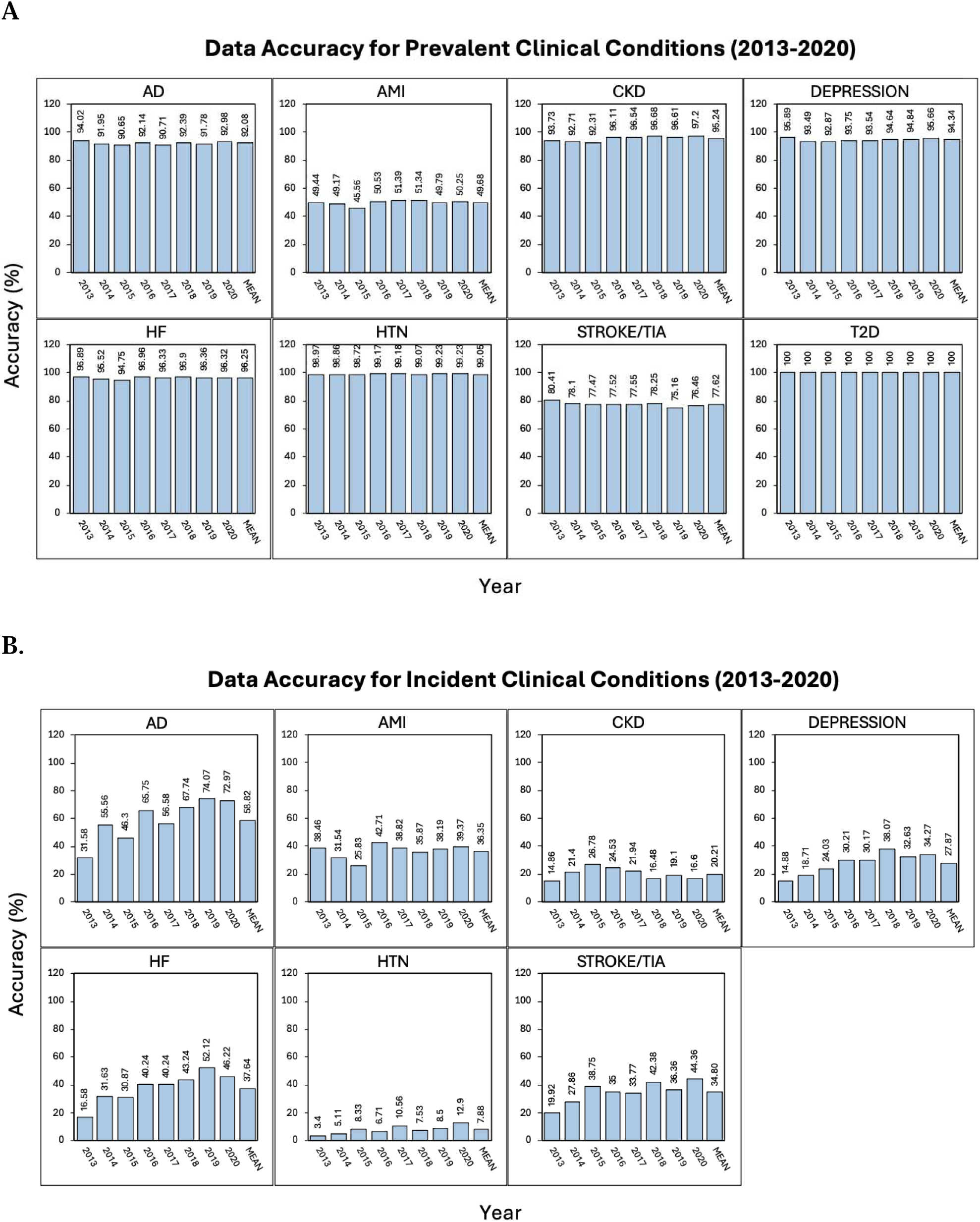
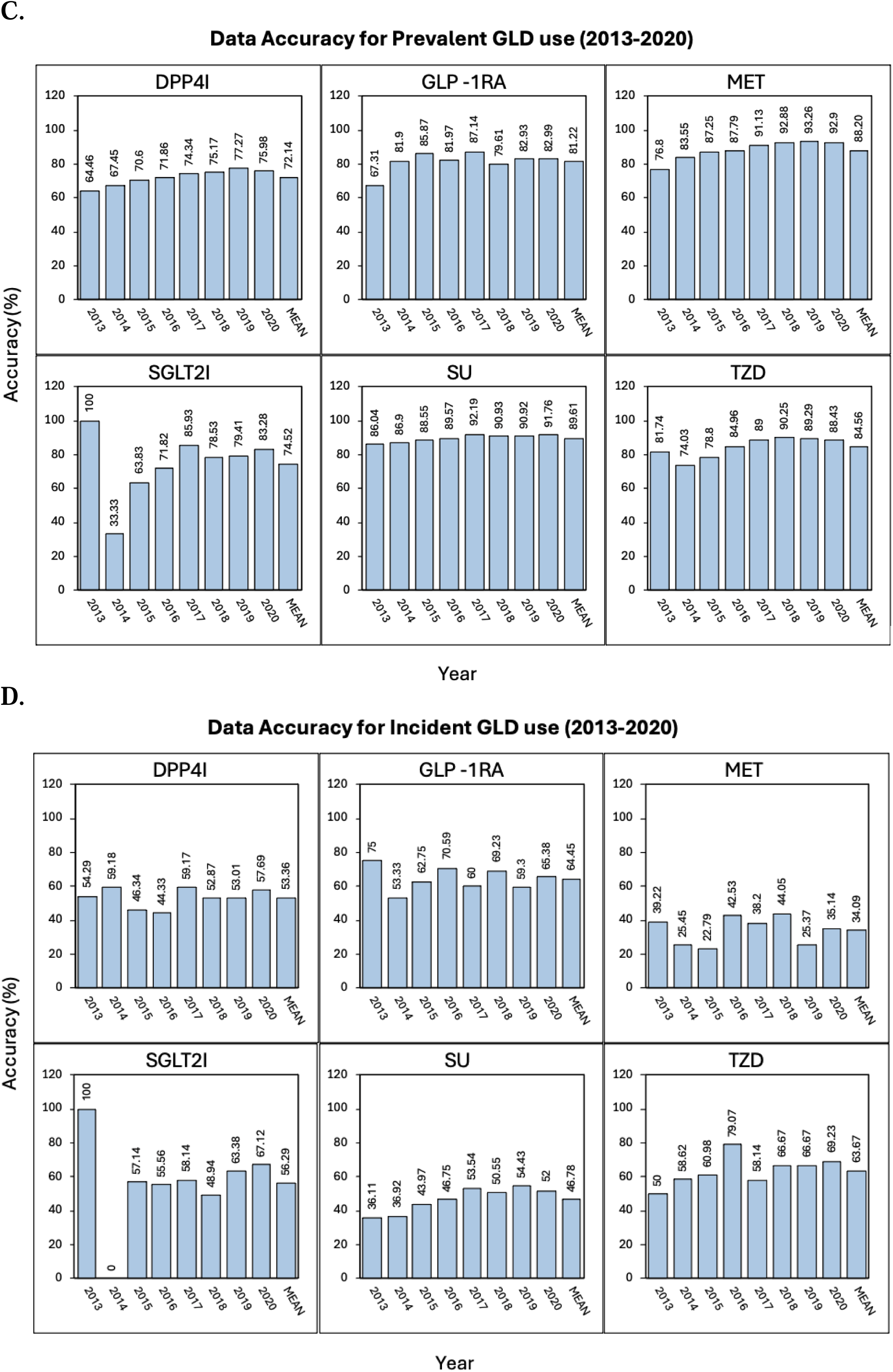
Accuracy of UF Health EHR data compared to Medicare claims for prevalent clinical conditions (A), incident clinical conditions (B), prevalent GLD use (C), and incident GLD use (D) in the UF Health EHR-Medicare linked T2D cohort (2013–2020). The proportion of accuracy values was calculated as: ***Accuracy rate = Number of patients whose EHR information matched Medicare / Number of patients with the element captured in EHR***

**Figure 3.**
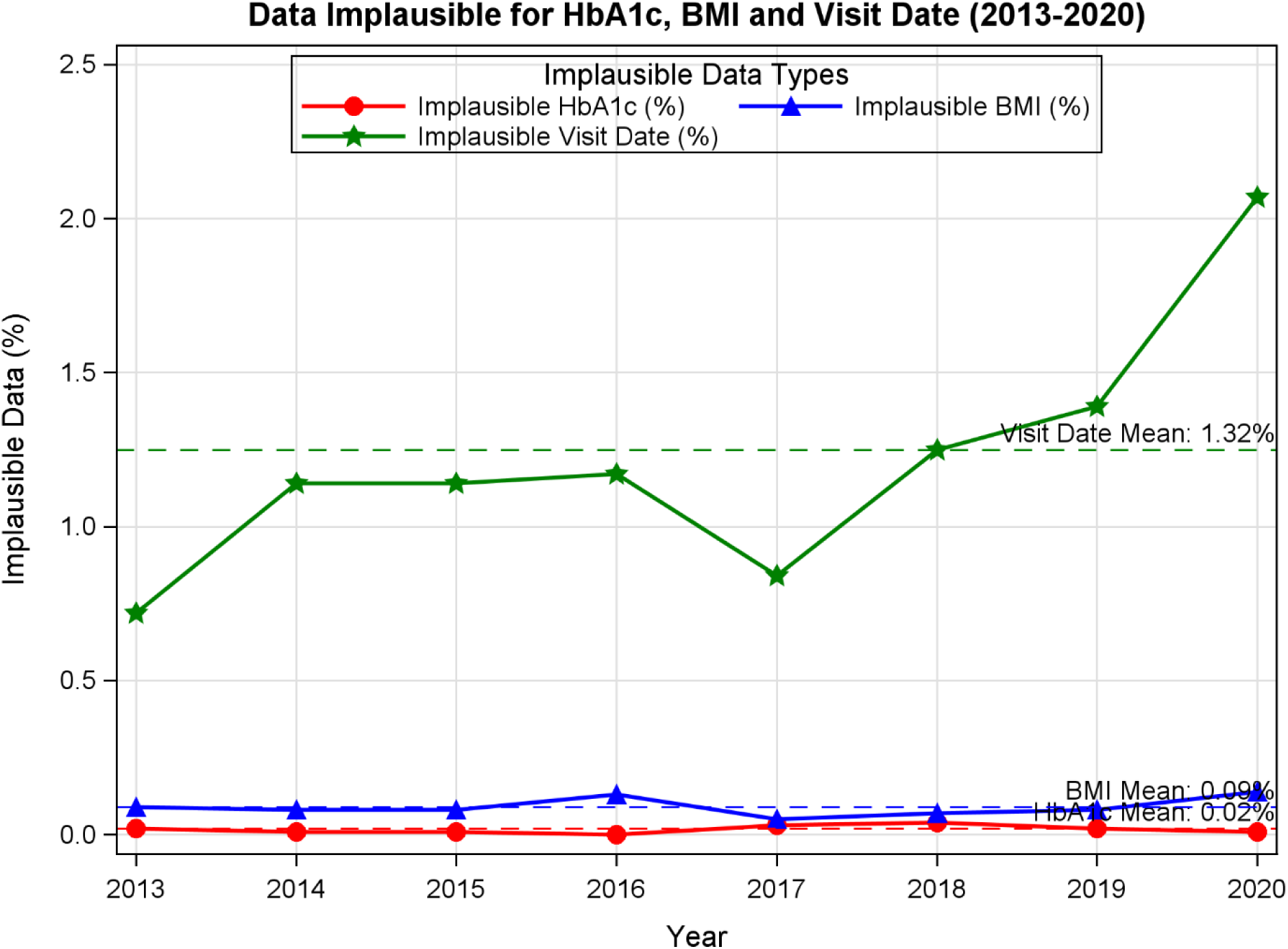
Plausibility of UF Health EHR data compared to Medicare claims for HbA1c, BMI, and visit date in the UF Health EHR-Medicare linked T2D cohort (2013–2020). The proportion of plausibility values was calculated as: ***Plausibility rate = Number of plausible values in EHR / Total number of values assessed in EHR***

Regarding medication data, the accuracy of prevalent GLD use varied by drug class (**Figure 2-C**). Traditional agents like SUs (89.6% accuracy, SD: 2.27), MET (88.2%, SD: 5.74), and TZD (84.6%, SD: 5.88) showed relatively good alignment with Medicare data. Newer agents such as GLP-1RA (71.2%, SD: 6.09), SGLT2i (74.5%, SD: 19.69), and DPP4i (72.1%, SD: 4.45) had lower accuracy rates. Documentation of incident GLD use was substantially less accurate across all classes (**Figure 2-D**), with MET showing the lowest accuracy (34.1%, SD: 8.39), followed by SU (46.8%, SD: 7.22), DPP4i (53.4%, SD: 5.59), SGLT2i (56.3%, SD: 27.53), TZD (63.7%, SD: 8.75), and GLP-1RA (64.4%, SD: 7.03), underscoring limitations in capturing the initiation of therapies in the EHR.

#### Plausibility of EHR data

The plausibility of key clinical values was generally high throughout the study period (**Table 3**). Plausible values were high for HbA1c (99.98%, SD: 0.01), BMI (99.91%, SD: 0.03), and visit dates (98.68%, SD: 0.48).

**Table 3.** Plausibility of UF Health EHR data in the UF Health EHR-Medicare linked T2D cohort (2013-2020)

| Characteristic | 2013 | 2014 | 2015 | 2016 | 2017 | 2018 | 2019 | 2020 | Mean (SD) |
| --- | --- | --- | --- | --- | --- | --- | --- | --- | --- |
| Cohort Size (n) | 10,354 | 12,025 | 13,251 | 13,990 | 14,846 | 14,243 | 12,525 | 11,922 | 12,895 (1476) |
| Plausible HbA1c (%) | 99.08 | 99.09 | 99.09 | 100 | 99.07 | 99.06 | 99.08 | 99.09 | 99.08 (0.01) |
| Plausible BMI (%) | 99.01 | 99.02 | 99.02 | 99.87 | 99.05 | 99.03 | 99.02 | 99.86 | 99.01 (0.03) |
| Plausible Visit Date (%) | 99.28 | 98.86 | 98.86 | 98.83 | 99.16 | 98.75 | 98.61 | 97.93 | 98.68 (0.48) |

Note: Plausible values were defined as follows — HbA1c: >2.5% and <25%; BMI: >12 and <70 kg/m<sup>2</sup>; Visit date: before birth or after death.

#### Concordance of EHR data

The concordance between EHR-documented HbA1c testing and T2D diagnosis remained consistently high throughout the study period (**Table 4**). Among patients with at least one HbA1c test result ≥6.5%, an average of 98.6% (SD: 0.25) also recorded a corresponding T2D diagnosis in the EHR. This high and stable concordance suggests strong internal consistency between laboratory results and diagnostic documentation for T2D in the UF Health EHR data across the years. This high level of internal consistency suggests that, within this healthcare system, laboratory-confirmed T2D diagnosis is reliably reflected in diagnostic coding practices.

**Table 4.**
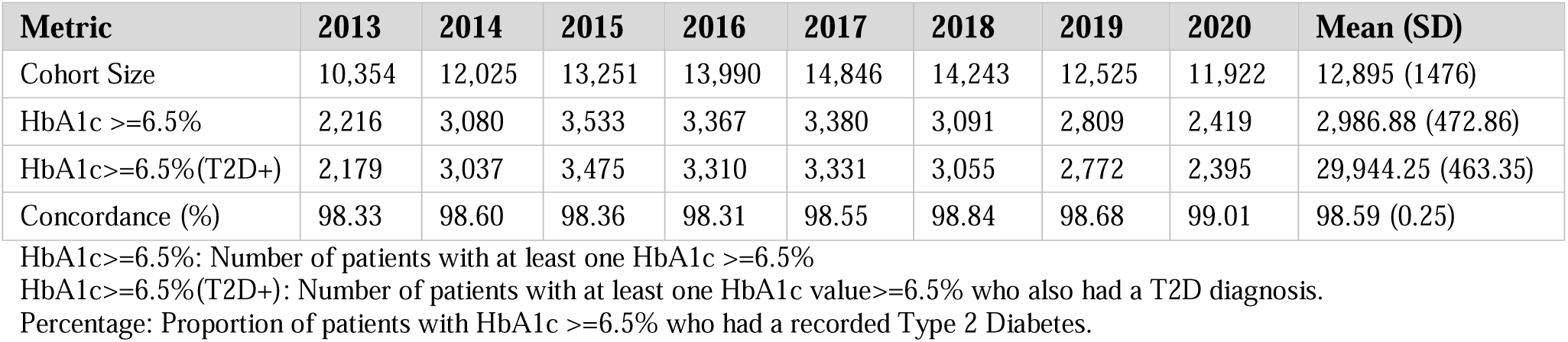
Concordance of UF Health EHR data between HbA1c testing and T2D diagnosis in the UF Health EHR-Medicare linked T2D cohort (2013-2020)

### Comparisons of data quality among EHR-only, Claims-only, and EHR-Claims linked data

In assessing data completeness, the UF Health EHR alone exhibited lower completeness compared to Medicare claims, particularly for mortality (68.2%), encounters (38–42%), and several clinical conditions (ranging from 34.1% to 78.4%). Laboratory results (e.g., HbA1c at 53.6%) and vital signs (e.g., BMI at 82.4%) were only available in the EHR, these data were not captured in Medicare. After linking EHR and claims data, the EHR-Claims linked data had complete capture (100%) across all demographics, mortality, encounter, clinical condition, and medication variables, while also incorporating granular clinical information from the EHR, such as BMI and HbA1c documentation **(Table S5).** In terms of data accuracy, the EHR alone demonstrated variable accuracies, with relatively high accuracy rates for demographic and prevalent clinical conditions (e.g., T2D: 100%, HTN: 99.05%) but low accuracy for incident clinical conditions (e.g., HTN: 7.9%, CKD: 20.2%) and incident GLD use (ranging from 34.1% to 64.4%). In contrast, no discordance were observed in the Medicare (reference) data or the linked EHR–claims cohort **(Table S6)**. These findings underscore the complementary strengths of Medicare and EHR data sources and highlight the substantial improvement in data completeness, accuracy, and clinical granularity through linkage.

### Representativeness of EHR-Medicare linked data

**Table S7** compares the demographic and clinical profiles of EHR data in the EHR-Medicare linked cohort with the national Medicare population with T2D. **Table S8** shows the differences by reporting the SMD and p-values. The EHR- Medicare linked cohort was slightly younger (mean age: 74.9 vs. 76. 5 years, SMD: -5.68, p<0.001) and had a higher proportion of females (58.0% vs. 56.6%, SMD: 0.03, p<0.001). Racial and ethnic distributions varied substantially; the EHR- Medicare linked group had more NHB (25.6% vs. 10.9%, SMD: 0.39, p<0.001) and fewer NHW (64.9% vs. 73.0%, SMD: -0.18, p<0.001) and Hispanic patients (3.6% vs. 10.2%, SMD: -0.27, p<0.001).

In terms of clinical characteristics, the EHR-Medicare linked T2D cohort had lower annual mortality (5.2% vs. 6.6%, SMD: -0.06, p<0.001) and HbA1c testing rate (37.8% vs. 72.9%, SMD: -0.76, p<0.001), indicating under-capture of routine diabetes monitoring. The EHR system identified only 65.8% of the T2D patients found in Medicare claims (SMD: -1.02, p<0.001). Prevalence of common comorbidities was consistently lower in the EHR cohort, including HTN (76.1% vs. 94.7%, SMD: -0.55), CKD (27.4% vs. 53.4%, SMD: -0.55), HF (21.6% vs. 39.2%, SMD: -0.39), depression (22.1% vs. 40.5%, SMD: -0.41), stroke/TIA (13.8% vs. 21.1%, SMD: -0.19), AMI (5.5% vs. 7.5%, SMD: -0.08), and AD (2.6% vs. 9.2%, SMD: -0.28) (all p<0.001). Despite the lower prevalence, the EHR-Medicare linked cohort had higher average incidence rates for several clinical conditions. Incidence of HTN (2.1% vs. 1.1%, SMD: 0.07), CKD (11.1% vs. 10.0%, SMD: 0.04), HF (13.8% vs. 7.4%, SMD: 0.21), depression (11.3% vs. 6.5%, SMD: 0.17), stroke/TIA (13.2% vs. 8.3%, SMD: 0.16), AMI (19.5% vs. 12.0%, SMD: 0.21), and AD (17.7% vs. 13.0%, SMD: 0.13) were significantly higher in the EHR group (all p<0.001), possibly reflecting differences in documentation or disease detection patterns.

Use of GLDs was consistently lower in the EHR-Medicare cohort across all classes. The largest difference was seen for MET (18.6% vs. 34.7%, SMD: -0.37, p<0.001). Other medications, including GLP-1RA (1.8% vs. 3.0%, SMD: -0.08), SGLT2i (1.1% vs. 2.3%, SMD: -0.10), DPP4i (4.9% vs. 7.6%, SMD: -0.11), SU (9.8% vs. 18.8%, SMD: -0.26), and TZD (1.8% vs. 3.2%, SMD: -0.09), were also underrepresented (all p<0.001). Incident GLD use showed similar trends, with lower uptake in the EHR group for most agents, except for SGLT2i, where use was comparable (37.2% vs. 37.6%, SMD: -0.01, p=0.41).

## Discussion

In this study, we evaluated the data quality of an EHR–Medicare linked cohort of older adults with T2D within a large academic health system, focusing on information critical for target trial emulation studies. Our findings indicate that while EHR data provides rich clinical detail, the completeness, accuracy, and representativeness relative to Medicare claims varied across domains and clinical variables. The linked EHR-Claims dataset demonstrated not only high quality but also contributed additional clinical details absent in claims, offering a more comprehensive and robust resource for real-world drug evaluation.

Demographic information—such as DOB, sex, and race/ethnicity—was complete and highly accurate, and BMI was well documented in the EHR, supporting the reliability of EHRs for capturing patient-level demographics. However, the EHR captured only about one-third of encounters, just over 50% of HbA1c tests, and nearly 70% of deaths recorded in Medicare, reflecting fragmented care among Medicare beneficiaries, who commonly receive services from multiple providers and across different health systems. As such, single-system EHRs may underestimate healthcare utilization and miss care delivered elsewhere. EHR data continuity has previously been recognized as a critical issue, and various strategies to measure and address it have been discussed in the literature[27–29]. In our recent work, we developed a data continuity algorithm to identify patients with high EHR data continuity, which is currently undergoing external validation. Additionally, as data shown in the current study, linking insurance claims data can also help capture external care, improving the completeness and granularity of EHR-based analyses.

Although prevalent clinical conditions were reasonably complete, the capture of incident (newly diagnosed) conditions was markedly poor across nearly all comorbidities examined. One possible explanation is the use of a one-year look-back period to define incident cases, which may have led to misclassification of some pre-existing conditions as new-onset. Extending the baseline period may improve the accuracy of incident condition identification. Additionally, integrating external data sources, such as Medicare claims, could offer longitudinal information that extends beyond the EHR’s data capture window, allowing more accurate distinction between prevalent and incident diagnoses.

The completeness of GLD documentation was limited, with most drug classes captured less than 60% of those documented in Medicare claims. The documentation of prevalent GLD use was more accurate for older agents such as MET and SUs, while newer agents including GLP-1RAs, SGLT2is, and DPP4is showed higher error rates. Accuracy was particularly poor for incident use, suggesting limited ability of EHRs to capture therapy initiation, which may be caused by the following reasons. First, a short one-year baseline period may fail to capture earlier medication exposures, misclassifying prevalent users as new users. Extending the baseline period may improve the accuracy of identifying incident medication by capturing prior exposures more comprehensively. Second, discrepancies between prescribing and dispensing, incomplete medication reconciliation, and system-level limitations in medication documentation may also contribute to under-capture. Given the increasing emphasis on evaluating uptake, effectiveness, and safety of newer GLDs, these limitations could bias pharmacoepidemiologic studies if not properly addressed. Linking EHR data with external sources such as pharmacy claims or Medicare Part D can enhance completeness and validity, particularly for prescriptions initiated outside the health system. Furthermore, such linkage enables capture of dispensing events and longitudinal medication adherence patterns, offering a more accurate reflection of actual drug exposure than EHR prescribing records alone. Additionally, employing advanced algorithms that leverage longitudinal prescription patterns and clinical notes may better identify true therapy initiation and changes.

Our analysis of data plausibility and concordance found that most clinical measures, such as HbA1c and BMI, were biologically plausible, with high rates of plausible values. In addition, we observed a high concordance between elevated HbA1c value and documented T2D diagnosis within the UF Health EHR. Together, the strong plausibility and internal concordance support the validity of using these EHR data elements for research, clinical quality assessment, and population health monitoring, with reduced concern that data artifacts or extreme outliers would meaningfully distort analyses. This plausibility-focused evaluation is consistent with established EHR data-quality frameworks and operational QA practices that assess reasonable value distributions and unit consistency[30]. These findings also highlight the value of routinely incorporating plausibility checks for frequently measured clinical variables and suggest that similar evaluations could be extended to other populations and disease areas by assessing condition-relevant laboratory measures and vital signs[31].

In interpreting our findings on the representativeness of the UF Health EHR–Medicare linked dataset relative to the national Medicare T2D population, we note meaningful differences in race/ethnicity composition and in capture of comorbidities, HbA1c testing, and GLD use. These differences may limit the transportability of our quantitative EHR-only data-quality estimates, as greater out-of-system care and setting-specific documentation patterns could reduce apparent completeness and alter diagnosis timing compared with more integrated settings. Therefore, our findings are most generalizable to similar academic health systems and patient mixes, while the overarching conclusion that Medicare linkage improves cross-setting capture of target trial emulation-relevant elements remains broadly applicable.

Overall, this study provides a novel and comprehensive evaluation of EHR data quality and representativeness by linking UF Health EHRs with Medicare claims for older adults with T2D. It adopted a structured framework to assess four key dimensions of data quality: completeness, accuracy, plausibility, and concordance, using Medicare claims as a reference. This multi-faceted approach offers a more rigorous evaluation than typically seen in prior EHR-based studies[32,33]. Importantly, the study distinguishes between prevalent and incident diagnoses and treatments, revealing limitations in the EHR’s ability to capture new disease events and medication initiations—critical for studies of treatment effects and disease progression. The longitudinal design, covering data from 2013 to 2020, allows for the assessment of data quality trends over time. Furthermore, by comparing the EHR–Medicare linked cohort to the national Medicare T2D population, we provide important context for the representativeness and transportability of single-site EHR findings.

These findings also have important implications for study designs of target trial emulations using RWD, including analyses that use new-user definitions, incident outcomes, and covariate adjustment. EHR-only gaps may introduce bias through misclassification of treatment initiation, under-ascertainment of incident outcomes, and incomplete capture of baseline covariates needed for confounding control. Incomplete encounter data can also complicate follow-up and censoring definitions. Linkage to claims mitigates several of these limitations while EHRs retain richer clinical detail.

Several limitations should be acknowledged. First, our analysis was limited to a single academic health system, and findings may not generalize to community-based or multi-institutional settings. In addition, analyses were restricted to older patients with T2D; thus, conclusions regarding the overall quality and added clinical value of the linked EHR–claims dataset should be interpreted within this population. Moreover, linkage was performed at the person level rather than the encounter/visit level, which precluded visit-level concordance analyses (e.g., assessing whether diagnoses occurred on the same visit across EHR and claims) and limited our ability to distinguish true out-of-system care from discrepancies driven by differences in how encounters are defined and recorded in EHR versus claims data. Second, we were unable to assess data quality for certain clinical domains (e.g., lifestyle factors, symptom burden) that are poorly captured in both EHRs and claims. Third, our definition of incident conditions and medication use relied on the absence of documentation in a prior look-back period, which may be sensitive to data completeness and follow-up duration. Finally, despite using Medicare claims as a gold standard, claims themselves may contain coding inaccuracies, though they are widely used for benchmarking in health services research.

Future work can address these limitations in several ways. First, generalizability can be strengthened by replicating the evaluation across additional health systems and/or multi-site networks and by validating key phenotypes across institutions[34,35]. Second, domains that are poorly captured in both EHR and claims (e.g., lifestyle factors and symptoms) may be supplemented using patient-reported outcomes, registries, or NLP-based extraction from clinical notes when available. Third, incident condition and initiation definitions can be evaluated under alternative washout/look-back windows (supported by continuous enrollment) and assessed with quantitative sensitivity/bias analyses to understand robustness to definitional choices[36]. Finally, because claims may contain coding errors, cross-source adjudication and routine plausibility/temporal data-quality checks can help identify inconsistencies prior to downstream analyses[30].

## Conclusions

While EHRs offer detailed clinical information, they present notable challenges in data quality assurance. Our study highlights the critical need to develop and implement robust data quality assessment strategies and frameworks when EHR data are used as a research source for target trial emulation studies. Linkage to external data sources such as claims, pharmacy records, or death registries can enhance data completeness and support robust observational studies. Future research should explore methods to improve EHR data quality through better integration of external care records, natural language processing, and standardized documentation workflows.

## Author contributions

Yao An Lee (Methodology, Writing—original draft), Ying Lu (Conceptualization, Data curation, Formal analysis, Methodology, Writing—original draft), Earl J Morris (Conceptualization, Investigation, Writing—review & editing), Xing He (Software, Validation, Resources), Yao An Lee (Software, Validation, Data curation), Aliment Winterstein (Supervision, Resources, Writing—review & editing), Jiang Bian (Conceptualization, Funding acquisition, Methodology, Writing—review & editing), and Jingchuan Guo (Supervision, Conceptualization, Funding acquisition, Methodology, Writing— review & editing).

## Data availability

The data used in this study are derived from linked UF Health EHRs and Medicare claims data. Due to privacy regulations and data use agreements, these data are not publicly available.

