## Supplemental Materials for "Assessing the Quality of Electronic Health Record Data and the Claims Linked Data for Target Trial Emulation Studies"

**Supplemental material**

**Contents**

**Table S1**. ICD-codes used for T2D and clinical conditions diagnosis

**Table S2.** Variables selection for quality assessment and their roles in target trial emulation (TTE) framework

**Table S3**. GLD list

**Table S4**. Demographics of UF Health EHR-Medicare claims linked T2D cohort (2013-2020)

**Table S5.** Comparison of Medicare, UF Health EHR and EHR-Medicare linked cohort for completeness

**Table S6.** Comparison of Medicare, UF Health EHR and EHR-Medicare linked cohort for inaccuracy

**Table S7**. Demographic and clinical characteristics of UF Health EHR data in EHR-Medicare linked cohort compared to National Medicare claims T2D population (2013 - 2020)

**Table S8.** Representativeness of UF Health EHR data in EHR-Medicare linked cohort compared to national T2D patients in Medicare claims

**Figure S1.** Flow Diagram of the UF Health–Medicare Linkage Process

**Table S1**. ICD-codes used for T2D and clinical conditions diagnosis

| **Clinical conditions** | **ICD-9** | **ICD-10** | **Period** | **CCW setting** |
| --- | --- | --- | --- | --- |
| T2D | 250.00, 250.2, 250.10, 250.12, 250.20, 250.22, 250.30, 250.32, 250.40, 250.42, 250.50, 250.52, 250.60, 250.62, 250.62, 250.70, 250.72, 250.80, 250.90, 250.92 | E11 | 2 years | At least 1 inpatient/SNF/HHA claim OR 2 HOP/carrier claims with DX codes |
| T1D | 250.01, 250.03, 250.11, 250.13, 250.21, 250.23, 250.31, 250.33, 250.41, 250.43, 250.51, 250.53, 250.61, 250.63, 250.71, 250.73, 250.81, 250.83, 250.91, 250.93 | E10 | 2 years | At least 1 inpatient/SNF/HHA claim OR 2 HOP/carrier claims with DX codes |
| CKD | 585 | N18 | 2 years | At least 1 inpatient/SNF/HHA claim OR 2 HOP/carrier claims with DX codes |
| HF | 428 | I50 | 2 years | At least 1 inpatient/SNF/HHA claim OR 2 HOP/carrier claims with DX codes |
| HTN | 401, 405 | I10, I15 | 2 years | At least 1 inpatient/SNF/HHA claim OR 2 HOP/carrier claims with DX codes |
| Depression | 296.2, 296.3, 296.51, 296.52, 296.53, 296.54, 296.55, 296.56, 296.6, 296.89, 298.0, 300.4, 309.1, 311 | F32, F33 | 2 years | At least 1 inpatient/SNF/HHA claim OR 2 HOP/carrier claims with DX codes |
| Stroke / TIA | 430, 431, 433.01, 433.11, 433.21, 433.31, 433.81, 433.91, 434.00, 434.01, 434.10, 434.11, 434.90, 434.91, 435.0, 435.1, 435.3, 435.8, 435.9, 436, 997.02 | G45.0, G45.1, G45.2, G45.3, G45.8, G45.9, G46, G97.3, I60, I61, I62.00, I62.02, I62.9, I63, I67.84, I67.89, I97.81, I97.82 | 1 year | At least 1 inpatient, HOP or carrier claim with DX codes |
| AD | 331.0 | G30.0 | 2 years | At least 1 inpatient/SNF/HHA claim OR 2 HOP/carrier claims with DX codes |
| AMI | 410 | I21, I22 | 1 year | At least 1 inpatient claim with DX codes |

**Table S2.** Variables selection for quality assessment and their roles in target trial emulation (TTE) framework

| **Target Trial Emulation framework** | **Information needed** | **Variables assessed in the current study** |
| --- | --- | --- |
| 1. Eligibility criteria | Baseline demographics, clinical conditions and medication use | Variables documenting demographics, prevalent clinical conditions and prevalent medication use |
| 2. Treatment strategies | Treatment of interest and comparators | Incident use of treatment with 1-year look-back (no record in prior year) |
| 3. Assignment procedures | Baseline information needed for computing propensity score | Same as No.1 |
| 4. Follow-up period | Encounter and mortality information | Encounter capture; mortality |
| 5. Outcome | Incidence of outcome of interest | Incident clinical condition |
| 6. Causal contrasts of interest | Intent to treat vs. per protocol analysis | Not applicable |
| 7. Analysis plan | Statistical model specification | Not applicable |

**Table S3**. GLD list

| **Category** | **Medications** |
| --- | --- |
| MET | metformin |
| GLP-1RA | Albiglutide*, Dulaglutide, Exenatide, Liraglutide, Lixisenatide, Semaglutide |
| SGLT2i | Canagliflozin, Dapagliflozin, Empagliflozin, Ertugliflozin |
| DPP4i | Alogliptin, Linagliptin, Saxagliptin, Sitagliptin |
| SU | Acetohexamide, Carbutamide, Chlorporpamide, Glibornuride, Glimepiride, Glipizide, Gliquidone, Glyburide, Tolazamide, Tolbutamide |
| TZD | Pioglitazone, Rosiglitazone |

*Withdraw from the market in 2018. GLP-1RAs, glucagon-like peptide-1 receptor agonists; SGLT2i, sodium-glucose cotransporter 2 inhibitors; DPP4i, dipeptidyl peptidase 4 inhibitors.

**Table S4**. Demographics of UF Health EHR-Medicare claims linked T2D cohort (2013-2020)

| **Characteristic** | **2013** | | **2014** | | **2015** | | **2016** | | **2017** | | **2018** | | **2019** | | **2020** | | **Mean** | |
| --- | --- | --- | --- | --- | --- | --- | --- | --- | --- | --- | --- | --- | --- | --- | --- | --- | --- | --- |
|  | **EHR** | **Medicare** | **EHR** | **Medicare** | **EHR** | **Medicare** | **EHR** | **Medicare** | **EHR** | **Medicare** | **EHR** | **Medicare** | **EHR** | **Medicare** | **EHR** | **Medicare** | **EHR** | **Medicare** |
| Number of patients | 10,354 | | 12,025 | | 13,251 | | 13,990 | | 14,846 | | 14,243 | | 12,525 | | 11,922 | | 12,895  (1476) | 12,895  (1476) |
| Age, mean (SD) | 74.37 (7.01) | 74.38  (7.02) | 74.53 (7.08) | 74.54  (7.07) | 74.67 (7.08) | 74.67  (7.07) | 74.72 (7.01) | 74.72  (7.07) | 74.81 (7.06) | 74.82  (7.06) | 75.06 (7.06) | 75.07  (7.06) | 75.21 (7.00) | 75.21  (7.01) | 75.51 (6.96) | 75.52  (6.96) | 74.86  (0.38) | 74.87  (0.38) |
| Female, n (%) | 6,259 (60.45) | 6259  (60.45) | 7,247 (60.27) | 7247  (60.27) | 7,933 (59.87) | 7933  (59.87) | 8,087 (57.74) | 8,078  (57.74) | 8,457 (56.96) | 8,457  (56.96) | 8,078 (56.72) | 8,078  (56.72) | 7,070 (56.45) | 7,070  (56.45) | 6,650 (55.78) | 6,650  (55.78) | 7473 (58.03) | 7472  (58.03) |
| **Race/ Ethnicity, n (%)** | | | | | | | | | | | | | | | | | | |
| NHW | 6,136 (59.26) | 6398  (61.79) | 7,261 (60.38) | 7489  (62.28) | 8,118 (61.26) | 8398  (63.38) | 9,147 (65.38) | 9,466  (67.66) | 9,847 (66.33) | 10,174(68.53) | 9,693 (68.05) | 9,930  (69.72) | 8,561 (68.35) | 8,726  (69.67) | 8,347 (70.01) | 8,538  (71.62) | 8389, (64.88) | 7707  (66.83) |
| NHB | 3,220 (31.10) | 3321  (32.07) | 3,648  (30.34) | 3738  (31.09) | 3,856 (29.10) | 3941  (29.74) | 3,459 (24.72) | 3,570  (25.52) | 3,502 (23.59) | 3,592  (24.20) | 3,186 (22.37) | 3,261  (22.90) | 2,802 (22.37) | 2,869  (22.91) | 2,526 (21.19) | 2,574  (21.59) | 32758  (25.60) | 3897 (26.25) |
| Hispanic | 346 (3.34) | 400  (3.86) | 425 (3.53) | 489  (4.07) | 499 (3.77) | 583  (4.40) | 501 (3.58) | 577  (4.12) | 573 (3.86) | 651  (4.39) | 515 (3.62) | 604  (4.24) | 441 (3.52) | 502  (4.01) | 377 (3.16) | 421  (3.53) | 460  (3.55) | 449  (4.08) |
| Other | 371 (3.58) | 214  (2.07) | 394 (3.28) | 258  (2.15) | 463 (3.49) | 272  (2.05) | 510 (3.65) | 296  (2.12) | 533 (3.59) | 324  (2.18) | 525 (3.69) | 324  (2.27) | 471 (3.76) | 302  (2.41) | 451 (3.78) | 269  (2.26) | 465  (3.6) | 239  (2.19) |
| Unknown | 281  (2.71) | 21  (0.20) | 297  (2.47) | 51  (0.42) | 315  (2.38) | 57  (0.43) | 373  (2.67) | 81  (0.58) | 391  (2.63) | 105  (0.71) | 324  (2.27) | 124  (0.87) | 250(2.00) | 126  (1.01) | 221  (1.85) | 120  (1.01) | 307  (2.37() | 72  (0.65) |

**Table S5.** Comparison of Medicare, UF Health EHR and EHR-Medicare linked cohort for completeness

| **Variable** | **Medicare** | **EHR** | **EHR-Medicare linked** |
| --- | --- | --- | --- |
| **Demographics (%)** | | | |
| DOB | 100.00 (0.00) | 100.00 (0.00) | 100.00 (0.00) |
| Sex | 100.00 (0.00) | 100.00 (0.00) | 100.00 (0.00) |
| Race / Ethnicity | 100.00 (0.00) | 100.00 (0.00) | 100.00 (0.00) |
| **Mortality (%)** | | | |
| Date of Death | 100.00 (0.00) | 68.22 (6.51) | 100.00 (0.00) |
| **Encounter (%)** | | | |
| Overall Encounter | 100.00 (0.00) | 38.34 (2.56) | 100.00 (0.00) |
| Inpatient Encounter | 100.00 (0.00) | 41.76 (5.00) | 100.00 (0.00) |
| Outpatient Encounter | 100.00 (0.00) | 37.32 (2.48) | 100.00 (0.00) |
| **Lab test (%)** | | | |
| HbA1c Test | - | 53.63 (4.53) | 53.63 (4.53) |
| **Vital sign (%)** | | | |
| BMI | - | 82.35 (2.42) | 82.35 (2.42) |
| **Clinical conditions (%)** | | | |
| CKD | 100.00 (0.00) | 43.00 (2.92) | 100.00 (0.00) |
| HF | 100.00 (0.00) | 44.69 (8.32) | 100.00 (0.00) |
| HTN | 100.00 (0.00) | 78.37 (5.09) | 100.00 (0.00) |
| T2D | 100.00 (0.00) | 65.81 (2.90) | 100.00 (0.00) |
| Depression | 100.00 (0.00) | 44.25 (7.34) | 100.00 (0.00) |
| Stroke/TIA | 100.00 (0.00) | 51.71 (7.95) | 100.00 (0.00) |
| AMI | 100.00 (0.00) | 63.82 (16.67) | 100.00 (0.00) |
| AD | 100.00 (0.00) | 34.13 (7.01) | 100.00 (0.00) |
| **GLD use (%)** | | | |
| MET | 100.00 (0.00) | 56.53 (7.14) | 100.00 (0.00) |
| GLP-1RA | 100.00 (0.00) | 57.20 (9.16) | 100.00 (0.00) |
| SGLT2i | 100.00 (0.00) | 53.65 (27.71) | 100.00 (0.00) |
| DPP4i | 100.00 (0.00) | 63.38 (6.65) | 100.00 (0.00) |
| SU | 100.00 (0.00) | 54.64 (7.07) | 100.00 (0.00) |
| TZD | 100.00 (0.00) | 58.01 (6.22) | 100.00 (0.00) |

**Table S6.** Comparison of Medicare, UF Health EHR and EHR-Medicare linked cohort for accuracy

| **Variable** | **Medicare** | **EHR** | **EHR-Medicare linked** |
| --- | --- | --- | --- |
| **Demographics (%)** | | | |
| DOB | 100.00(0.00) | 99.31(0.20) | 100.00(0.00) |
| Sex | 100.00(0.00) | 100.00(0.00) | 100.00(0.00) |
| Race / Ethnicity | 100.00(0.00) | 92.58(0.20) | 100.00(0.00) |
| **Mortality (%)** | | | |
| Date of Death | 100.00(0.00) | 96.73(0.85) | 100.00(0.00) |
| **Prevalent chronic condition prevalence (%)** | | | |
| CKD | 100.00(0.00) | 95.24(1.98) | 100.00(0.00) |
| HF | 100.00(0.00) | 96.25(0.77) | 100.00(0.00) |
| HTN | 100.00(0.00) | 99.05(0.19) | 100.00(0.00) |
| T2D | 100.00(0.00) | 100.00 (0.00) | 100.00(0.00) |
| Depression | 100.00(0.00) | 94.33(1.09) | 100.00(0.00) |
| Stroke/TIA | 100.00(0.00) | 77.61(1.5) | 100.00(0.00) |
| AMI | 100.00(0.00) | 49.68(1.85) | 100.00(0.00) |
| AD | 100.00(0.00) | 92.08(1.11) | 100.00(0.00) |
| **Incident clinical condition (%)** | | | |
| CKD | 100.00(0.00) | 20.21(4.19) | 100.00(0.00) |
| HF | 100.00(0.00) | 37.64(11.04) | 100.00(0.00) |
| HTN | 100.00(0.00) | 7.88(2.98) | 100.00(0.00) |
| Depression | 100.00(0.00) | 27.87(7.98) | 100.00(0.00) |
| Stroke/TIA | 100.00(0.00) | 34.80(7.91) | 100.00(0.00) |
| AMI | 100.00(0.00) | 36.35(5.31) | 100.00(0.00) |
| AD | 100.00(0.00) | 58.82(14.51) | 100.00(0.00) |
| **Prevalent GLD use (%)** | | | |
| MET | 100.00(0.00) | 88.19(5.74) | 100.00(0.00) |
| GLP-1RA | 100.00(0.00) | 81.21(6.09) | 100.00(0.00) |
| SGLT2i | 100.00(0.00) | 74.52(19.69) | 100.00(0.00) |
| DPP4i | 100.00(0.00) | 72.14(4.45) | 100.00(0.00) |
| SU | 100.00(0.00) | 89.61(2.27) | 100.00(0.00) |
| TZD | 100.00(0.00) | 84.56(5.88) | 100.00(0.00) |
| **Incident GLD use (%)** | | | |
| MET | 100.00(0.00) | 34.09(8.39) | 100.00(0.00) |
| GLP-1RA | 100.00(0.00) | 64.45(7.03) | 100.00(0.00) |
| SGLT2i | 100.00(0.00) | 56.28(27.53) | 100.00(0.00) |
| DPP4i | 100.00(0.00) | 53.36(5.59) | 100.00(0.00) |
| SU | 100.00(0.00) | 46.78(7.22) | 100.00(0.00) |
| TZD | 100.00(0.00) | 63.67(8.75) | 100.00(0.00) |

**Table S7**. Demographic and clinical characteristics of UF Health EHR data in EHR-Medicare linked cohort compared to National Medicare claims T2D population (2013 - 2020)

| **Characteristic** | **Cohort** | **2013** | **2014** | **2015** | **2016** | **2017** | **2018** | **2019** | **2020** | **Mean (SD)** |
| --- | --- | --- | --- | --- | --- | --- | --- | --- | --- | --- |
| **Number of patients** | Linked | 10,354 | 12,025 | 13,251 | 13,990 | 14,846 | 14,243 | 12,525 | 11,922 | 12,895  (1,476) |
|  | National Medicare | 753,719 | 789,438 | 787,433 | 1,460,775 | 1,533,326 | 1,468,143 | 1,386,471 | 1,386,419 | 1195716  (35019) |
| **Demographic characteristic** (%) | | | | | | | | | | |
| Age, mean (SD) | Linked | 74.37  (7.01) | 74.53  (7.08) | 74.67  (7.08) | 74.72  (7.07) | 74.81  (7.06) | 75.06  (7.06) | 75.21  (7.01) | 75.51  (6.96) | 74.86  (0.38) |
|  | National Medicare | 76.57  (7.79) | 76.60  (7.84) | 76.36  (7.77) | 76.32  (7.80) | 76.35  (7.82) | 76.41  (7.82) | 76.43  (7.81) | 76.54  (7.77) | 76.45  (0.11) |
| Female, n (%) | Linked | 6,259  (60.45) | 7,247  (60.27) | 7,933  (59.87) | 8,087  (57.74) | 8,457  (56.96) | 8,078  (56.72) | 7,070  (56.45) | 6,650  (55.78) | 7473  (58.03) |
|  | National Medicare | 457,963  (60.76) | 463,270  (58.68) | 430,773  (54.71) | 822,741  (56.32) | 859,856  (56.08) | 818,763  (55.77) | 766,567  (55.29) | 759,336  (54.77) | 672,409  (56.55) |
| **Race/Ethnicity** (%) | | | | | | | | | | |
| NHW | Linked | 59.26 | 60.38 | 61.26 | 65.38 | 66.33 | 68.05 | 68.35 | 70.01 | 64.88  (4.07) |
|  | National Medicare | 72.97 | 72.5 | 71.81 | 72.92 | 72.59 | 73.5 | 73.71 | 74.26 | 73.03  (0.77) |
| NHB | Linked | 31.1 | 30.34 | 29.1 | 24.72 | 23.59 | 22.37 | 22.37 | 21.19 | 25.6  (3.97) |
|  | National Medicare | 11.81 | 11.87 | 11.91 | 10.92 | 10.91 | 10.24 | 10.02 | 9.62 | 10.91  (0.9) |
| Hispanic | Linked | 3.34 | 3.53 | 3.77 | 3.58 | 3.86 | 3.62 | 3.52 | 3.16 | 3.55  (0.22) |
|  | National Medicare | 10.51 | 10.66 | 10.95 | 10.42 | 10.49 | 9.93 | 9.64 | 9.24 | 10.23  (0.57) |
| Other | Linked | 3.58 | 3.28 | 3.49 | 3.65 | 3.59 | 3.69 | 3.76 | 3.78 | 3.6  (0.16) |
|  | National Medicare | 4.28 | 4.41 | 4.58 | 4.75 | 4.86 | 4.98 | 5.11 | 5.18 | 4.77  (0.33) |
| Unknown | Linked | 2.71 | 2.47 | 2.38 | 2.67 | 2.63 | 2.27 | 2 | 1.85 | 2.37  (0.32) |
|  | National Medicare | 0.42 | 0.57 | 0.75 | 0.99 | 1.16 | 1.35 | 1.53 | 1.71 | 1.06  (0.46) |
| **Annual mortality (%)** | | | | | | | | | | |
| Date of Death | Linked | 4.27 | 4.92 | 4.98 | 3.91 | 4.71 | 5.64 | 6.10 | 7.00 | 5.19  (1.01) |
|  | National Medicare | 6.16 | 6.40 | 6.61 | 5.95 | 6.62 | 6.69 | 6.66 | 8.06 | 6.64  (0.63) |
| **Number of encounters** | | | | | | | | | | |
| **Total encounter** | Linked | 138,142 | 170,392 | 202,026 | 205,542 | 221,092 | 225,092 | 212,769 | 201,356 | 197052  (2901) |
|  | National Medicare | 20,580,348 | 20,979,385 | 20,248,209 | 44,656,074 | 45,658,789 | 46,290,185 | 44,334,281 | 40,720,411 | 3543346  (12391160) |
| **HbA1c Testing Rates** (%) | | | | | | | | | | |
| **Lab test** | Linked | 35.03 | 39.46 | 41.14 | 37.71 | 37.04 | 37.39 | 38.14 | 36.45 | 37.8  (1.86) |
|  | National Medicare | 69.74 | 67.6 | 65.02 | 77.86 | 74.67 | 77.41 | 77.21 | 74.03 | 72.94  (4.9) |
| **Prevalent chronic condition** (%) | | | | | | | | | | |
| T2D | Linked | 59.32 | 65.84 | 66.78 | 65.16 | 66.2 | 66.08 | 68.5 | 68.59 | 65.81  (2.9) |
|  | National Medicare | 100 | 100 | 100 | 100 | 100 | 100 | 100 | 100 | 100  (0) |
| CKD | Linked | 18.96 | 23.27 | 26.09 | 26.85 | 28.2 | 29.78 | 32.3 | 33.51 | 27.37  (4.75) |
|  | National Medicare | 39.3 | 40.66 | 43.74 | 54.65 | 58.68 | 61.51 | 63.59 | 65.39 | 53.44  (10.67) |
| HF | Linked | 13.37 | 18.01 | 20.14 | 20.93 | 22.6 | 23.98 | 26.35 | 27.34 | 21.59  (4.56) |
|  | National Medicare | 39.96 | 39.36 | 38.35 | 39.52 | 39.3 | 39.28 | 39.06 | 38.75 | 39.2  (0.49) |
| HTN | Linked | 65.8 | 74 | 75.79 | 75.33 | 76.44 | 78.11 | 80.77 | 82.16 | 76.05  (4.98) |
|  | National Medicare | 94.64 | 94.53 | 94.27 | 94.97 | 94.93 | 94.91 | 94.76 | 94.65 | 94.71  (0.24) |
| Depression | Linked | 13.41 | 18.4 | 20.84 | 21.14 | 23.13 | 24.64 | 27.1 | 28.01 | 22.08  (4.77) |
|  | National Medicare | 36.33 | 37.27 | 37.84 | 40.79 | 41.85 | 42.82 | 43.46 | 43.85 | 40.53  (2.98) |
| Stroke/TIA | Linked | 9.46 | 12.15 | 13.37 | 13.35 | 14.37 | 15.17 | 15.88 | 16.71 | 13.81  (2.3) |
|  | National Medicare | 21.2 | 21.07 | 20.62 | 21.38 | 21.32 | 21.28 | 21.08 | 20.92 | 21.11  (0.25) |
| AMI | Linked | 2.58 | 4.01 | 4.59 | 4.71 | 5.35 | 6.81 | 7.62 | 8.51 | 5.52  (1.98) |
|  | National Medicare | 7.29 | 7.22 | 7.11 | 7.47 | 7.56 | 7.7 | 7.76 | 7.78 | 7.49  (0.26) |
| AD | Linked | 1.78 | 2.48 | 2.34 | 2.64 | 2.75 | 2.77 | 2.91 | 2.99 | 2.58  (0.39) |
|  | National Medicare | 9.25 | 9.14 | 8.88 | 9.54 | 9.53 | 9.39 | 9.09 | 8.69 | 9.19  (0.3) |
| **Incident clinical condition** (%) | | | | | | | | | | |
| CKD | Linked | 20.22 | 15.87 | 12.21 | 8.57 | 9.36 | 8.44 | 7.12 | 6.63 | 11.05  (4.77) |
|  | National Medicare | 11.39 | 11.34 | 14.28 | 7.66 | 11.74 | 8.95 | 7.92 | 6.32 | 9.95  (2.66) |
| HF | Linked | 27.02 | 18.98 | 13.71 | 11.2 | 12.37 | 9.95 | 10 | 7.3 | 13.82  (6.35) |
|  | National Medicare | 7.28 | 7.64 | 7.93 | 3.4 | 8.55 | 8.46 | 8.52 | 7.22 | 7.38  (1.69) |
| HTN | Linked | 5.61 | 2.64 | 1.91 | 1.56 | 1.25 | 1.31 | 1.14 | 0.95 | 2.05  (1.54) |
|  | National Medicare | 1.31 | 1.25 | 1.23 | 0.51 | 1.29 | 1.2 | 1.16 | 0.98 | 1.12  (0.27) |
| Depression | Linked | 29.54 | 18.84 | 11.15 | 7.94 | 7.05 | 6.21 | 5.6 | 4.28 | 11.33  (8.67) |
|  | National Medicare | 7.66 | 7.6 | 7.55 | 3.08 | 7.38 | 6.88 | 6.62 | 5.25 | 6.5  (1.6) |
| Stroke/TIA | Linked | 25.1 | 19.16 | 13.55 | 11.78 | 10.68 | 9.72 | 8.85 | 6.68 | 13.19  (6.08) |
|  | National Medicare | 8.82 | 9.22 | 9.05 | 3.92 | 9.41 | 9.17 | 9.04 | 7.86 | 8.31  (1.84) |
| AMI | Linked | 29.21 | 26.97 | 19.74 | 14.57 | 19.14 | 18.97 | 15.09 | 12.51 | 19.53  (5.89) |
|  | National Medicare | 12.21 | 12.27 | 12.49 | 5.6 | 14.24 | 13.88 | 13.69 | 11.72 | 12.01  (2.75) |
| AD | Linked | 20.65 | 24.16 | 17.42 | 19.78 | 18.58 | 15.74 | 14.79 | 10.39 | 17.69  (4.16) |
|  | National Medicare | 13.92 | 14.21 | 15.4 | 6.47 | 15.26 | 13.88 | 13.22 | 11.23 | 12.95  (2.92) |
| **Prevalent GLD use** (%) | | | | | | | | | | |
| MET | Linked | 12.86 | 20.07 | 21.72 | 19.96 | 20.19 | 18.83 | 18.25 | 16.77 | 18.58  (2.74) |
|  | National Medicare | 26.6 | 28.57 | 30.03 | 37.84 | 38.6 | 38.41 | 38.67 | 38.44 | 34.65  (5.26) |
| GLP-1RA | Linked | 0.5 | 0.87 | 1.39 | 1.31 | 1.89 | 2.17 | 2.99 | 3.3 | 1.8  (0.98) |
|  | National Medicare | 1.27 | 1.42 | 1.71 | 2.51 | 3.02 | 3.7 | 4.6 | 5.37 | 2.95  (1.51) |
| SGLT2i | Linked | 0 | 0.3 | 0.71 | 0.79 | 0.91 | 1.34 | 1.9 | 2.41 | 1.05  (0.8) |
|  | National Medicare | 0.12 | 0.58 | 1.3 | 2.13 | 2.7 | 3.05 | 3.92 | 4.91 | 2.34  (1.64) |
| DPP4i | Linked | 3.21 | 4.93 | 5.49 | 5.41 | 5.85 | 5.29 | 4.85 | 4.5 | 4.94  (0.82) |
|  | National Medicare | 6.51 | 6.73 | 6.97 | 8.58 | 8.5 | 8.12 | 7.78 | 7.2 | 7.55  (0.81) |
| SU | Linked | 8.99 | 12.37 | 12.32 | 10.62 | 9.92 | 8.83 | 8.18 | 7.13 | 9.8  (1.89) |
|  | National Medicare | 18.95 | 18.27 | 17.7 | 20.59 | 19.92 | 19 | 18.2 | 17.44 | 18.76  (1.08) |
| TZD | Linked | 1.11 | 1.51 | 1.89 | 1.9 | 1.96 | 1.94 | 2.01 | 2.03 | 1.79  (0.32) |
|  | National Medicare | 2.76 | 2.62 | 2.71 | 3.37 | 3.43 | 3.5 | 3.6 | 3.67 | 3.21  (0.43) |
| **Incident clinical condition** | | | | | | | | | | |
| MET | Linked | 3.83 | 2.28 | 4.73 | 3.11 | 2.97 | 3.13 | 2.93 | 3.7 | 3.34  (0.74) |
|  | National Medicare | 8.85 | 10.87 | 8.5 | 2.99 | 7.37 | 6.53 | 6.74 | 5.5 | 7.17  (2.37) |
| GLP-1RA | Linked | 15.38 | 14.29 | 27.72 | 18.58 | 23.21 | 21.04 | 22.93 | 19.8 | 20.37  (4.38) |
|  | National Medicare | 24.14 | 23.97 | 25.94 | 9.61 | 26.55 | 26.59 | 25.95 | 20.98 | 22.97  (5.71) |
| SGLT2i | Linked | 100 | 16.67 | 44.68 | 24.55 | 31.85 | 24.61 | 29.83 | 25.44 | 37.2  (26.62) |
|  | National Medicare | 58.35 | 64.6 | 50.32 | 12.77 | 31.53 | 26.26 | 30.59 | 25.96 | 37.55  (18.08) |
| DPP4i | Linked | 10.54 | 8.26 | 16.9 | 12.81 | 13.81 | 11.55 | 13.67 | 14.53 | 12.76  (2.64) |
|  | National Medicare | 20.11 | 22.08 | 22.42 | 7.67 | 19.79 | 17.99 | 17.33 | 14.54 | 17.74  (4.82) |
| SU | Linked | 3.87 | 4.37 | 7.1 | 5.18 | 6.73 | 7.24 | 7.71 | 5.88 | 6.01  (1.42) |
|  | National Medicare | 8.08 | 9.34 | 9.67 | 3.77 | 10.12 | 9.68 | 9.76 | 9.13 | 8.69  (2.08) |
| TZD | Linked | 3.48 | 16.02 | 16.4 | 16.17 | 14.78 | 15.16 | 20.24 | 10.74 | 14.12  (5.02) |
|  | National Medicare | 11.18 | 15.4 | 18.31 | 6.81 | 16.83 | 16.76 | 17.03 | 15.04 | 14.67  (3.83) |

**Table S8.** Representativeness of UF Health EHR data in EHR-Medicare linked cohort compared to national T2D patients in Medicare claims

| **Characteristic** | **EHR-Medicare linked cohort** | **National Medicare cohort** | **Raw difference** | **SMD** | **P-value** |
| --- | --- | --- | --- | --- | --- |
| **Number of patients, mean (SD)** | 12,895 (1,476) | 1195716 (350168.64) | -1,182,821 | - | - |
| **Demographics, mean percentage (SD)** | | | | | |
| Age, mean (SD) | 74.86 (0.38) | 76.45 (0.11) | -1.59 | -5.68 | <.0001 |
| **Female, mean percentage (SD)** | 58.03 (1.88) | 56.55 (2.11) | 1.48 | 0.03 | 0.0006 |
| **Race/Ethnicity, mean percentage (SD)** | | | | | |
| NHW | 64.88 (4.07) | 73.03 (0.77) | -8.15 | -0.18 | <.0001 |
| NHB | 25.60 (3.97) | 10.91 (0.9) | 14.69 | 0.39 | <.0001 |
| Hispanic | 3.55 (0.22) | 10.23 (0.57) | -6.68 | -0.27 | <.0001 |
| Other | 3.60 (0.16) | 4.77 (0.33) | -1.17 | -0.06 | <.0001 |
| Unknown | 2.37 (0.32) | 1.06 (0.46) | 1.31 | 0.10 | <.0001 |
| **Mortality, mean percentage (SD)** | | | | | |
| Date of Death | 5.19 (1.01) | 6.64 (0.63) | -1.45 | -0.06 | <.0001 |
| **HbA1c testing, mean percentage (SD)** | 37.8 (1.86) | 72.94 (4.9) | -35.14 | -0.76 | <0.001 |
| **Prevalent clinical condition, mean percentage (SD)** | | | | | |
| T2D | 65.81 (2.9) | 100.00 (0.00) | -34.19 | -1.02 | <.0001 |
| HTN | 76.05 (4.98) | 94.71 (0.24) | -18.66 | -0.55 | <.0001 |
| CKD | 27.37 (4.75) | 53.44 (10.67) | -26.07 | -0.55 | <.0001 |
| HF | 21.59 (4.56) | 39.20 (0.49) | -17.61 | -0.39 | <.0001 |
| Depression | 22.08 (4.77) | 40.53 (2.98) | -18.45 | -0.41 | <.0001 |
| Stroke/TIA | 13.81 (2.3) | 21.11 (0.25) | -7.30 | -0.19 | <.0001 |
| AMI | 5.52 (1.98) | 7.49 (0.26) | -1.97 | -0.08 | <.0001 |
| AD | 2.58 (0.39) | 9.19 (0.3) | -6.61 | -0.28 | <.0001 |
| **Incident clinical condition, mean percentage (SD)** | | | | | |
| HTN | 2.05 (1.54) | 1.12 (0.27) | 0.93 | 0.07 | <.0001 |
| CKD | 11.05 (4.77) | 9.95 (2.66) | 1.10 | 0.04 | <.0001 |
| HF | 13.82 (6.35) | 7.38 (1.69) | 6.44 | 0.21 | <.0001 |
| Depression | 11.33 (8.67) | 6.50 (1.60) | 4.83 | 0.17 | <.0001 |
| Stroke/TIA | 13.19 (6.08) | 8.31 (1.84) | 4.88 | 0.16 | <.0001 |
| AMI | 19.53 (5.89) | 12.01 (2.75) | 7.52 | 0.21 | <.0001 |
| AD | 17.69 (4.16) | 12.95 (2.92) | 4.74 | 0.13 | <.0001 |
| **Prevalent GLD use, mean percentage (SD)** | | | | | |
| MET | 18.58 (2.74) | 34.65 (5.26) | -16.07 | -0.37 | <.0001 |
| GLP-1RA | 1.80 (0.98) | 2.95 (1.51) | -1.15 | -0.08 | <.0001 |
| SGLT2i | 1.05 (0.8) | 2.34 (1.64) | -1.29 | -0.10 | <.0001 |
| DPP4i | 4.94 (0.82) | 7.55 (0.81) | -2.61 | -0.11 | <.0001 |
| SU | 9.80 (1.89) | 18.76 (1.08) | -8.96 | -0.26 | <.0001 |
| TZD | 1.79 (0.32) | 3.21 (0.43) | -1.42 | -0.09 | <.0001 |
| **Incident GLD Use, mean percentage (SD)** | | | | | |
| MET | 3.34 (0.74) | 7.17 (2.37) | -3.83 | -0.17 | <.0001 |
| GLP-1RA | 20.37 (4.38) | 22.97 (5.71) | -2.60 | -0.06 | <.0001 |
| SGLT2i | 37.20(26.62) | 37.55 (18.08) | -0.35 | -0.01 | 0.41 |
| DPP4 | 12.76 (2.64) | 17.74 (4.82) | -4.98 | -0.14 | <.0001 |
| SU | 6.01 (1.42) | 8.69 (2.08) | -2.68 | -0.10 | <.0001 |
| TZD | 14.12 (5.02) | 14.67 (3.83) | -0.55 | -0.02 | <.0001 |

**Figure S1.** Flow Diagram of the UF Health–Medicare Linkage Process

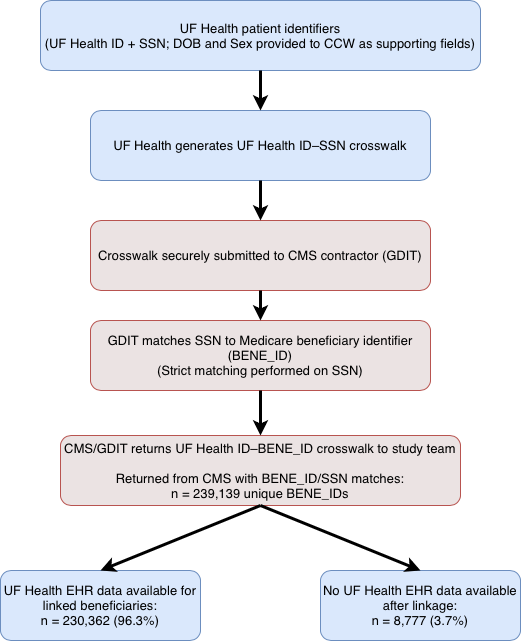

SSN: Social security number; DOB: Date of birth; CCW: Chronic Conditions Warehouse; CMS: Centers for Medicare & Medicaid Services; GDIT: General Dynamics Information Technology
